# High-resolution characterization of recent tuberculosis transmission in Botswana using geospatial and genomic data – the Kopanyo Study

**DOI:** 10.1101/2022.04.13.22273731

**Authors:** Chelsea R. Baker, Ivan Barilar, Leonardo S. de Araujo, Anne W. Rimoin, Daniel M. Parker, Rosanna Boyd, James L. Tobias, Patrick K. Moonan, Eleanor S. Click, Alyssa Finlay, John E. Oeltmann, Vladimir N. Minin, Chawangwa Modongo, Nicola M. Zetola, Stefan Niemann, Sanghyuk S. Shin

## Abstract

**Introduction:** Combining genomic and geospatial data can be useful for understanding *Mycobacterium tuberculosis* (Mtb) transmission in high tuberculosis burden settings.

**Methods:** We performed whole genome sequencing (WGS) on Mtb DNA extracted from sputum cultures from a population-based tuberculosis study conducted in 2012–2016 in Gaborone, Botswana. We used kernel density estimation, spatial *K*-functions, and created spatial distributions of phylogenetic trees. WGS-based clusters of isolates ≤5 single nucleotide polymorphisms were considered recent transmission, and large WGS-based clusters (≥10 members) were considered outbreaks.

**Results:** We analyzed data from 1449 participants with culture-confirmed TB. Among these, 946 (65%) participants had both molecular and geospatial data. A total of 62 belonged to five large outbreaks (10–19 participants each). Geospatial clustering was detected in two of the five large outbreaks, suggesting heterogeneous spatial patterns within the community.

**Conclusions:** Integration of genomic and geospatial data identified distinct patterns of tuberculosis transmission in a high-tuberculosis burden setting. Targeted interventions in these smaller geographies may interrupt on-going transmission.

## Introduction

Tuberculosis (TB) remains among the leading causes of infectious disease mortality worldwide, killing 1.5 million people in 2020 despite being a curable and preventable disease (1). High TB burden countries often contend with limited financial and labor resources, and typically rely on generalized interventions that, while important, treat TB as a uniform epidemic (2–4). However, recent advances in molecular methods have shown that TB epidemics are comprised of multiple simultaneous chains of transmission that could serve as distinct targets for intervention (4–7). Targeted interventions to interrupt transmission may be particularly effective for reducing TB in high-burden settings, where recent infection contributes substantially to disease incidence (3–6).

Genomic sequencing is a powerful tool for identifying closely related *M. tuberculosis* (Mtb) strains, helping to facilitate the reconstruction of likely chains of recent transmission (3,8,9). Genomic data can be integrated with geospatial data to investigate whether transmission chains are geographically distinct (3–6). For example, spatial clusters of closely related Mtb strains may indicate local areas of ongoing transmission, which could be targeted for public health interventions such as active case finding (3–5,10). A growing body of evidence suggests that geographically targeted interventions may be effective and cost-efficient in high-burden, low-resource settings, and could be instrumental in accelerating progress toward TB elimination (4,11–13).

In the Kopanyo Study, a population-based study of TB transmission in Botswana, localized transmission events were characterized by detecting spatial clustering of participants belonging to genotypic cluster groups identified using Mycobacterial Interspersed Repetitive Unit-Variable Number Tandem Repeat (MIRU-VNTR) genotyping (5,14). The objective of the current analysis was to build on the original study by incorporating higher resolution genomic data from whole genome sequencing (WGS), and to investigate the geographic distribution of distinct genotypic cluster groups representing potential recent transmission chains.

## Methods

### Study design and setting

We analyzed data collected for the Kopanyo Study among people with TB in Botswana from 2012–2016 (5,14). Botswana is a country in southern Africa with a high burden of TB and TB-HIV coinfection (1,5,14). Participants included men and women of all ages who were diagnosed with TB disease and sequentially enrolled in the study (5,14). Those who had already received TB treatment for 14 days or more, prisoners, and patients who declined to participate were excluded (5,14). At least one sputum sample was collected from each participant for bacterial culture (5,14). Clinical and demographic data, including residential address, were collected through in-person interviews and medical record review (5,14). Geographic coordinates for residential locations were obtained either by using by global positioning system (GPS) devices during site visits, or by geocoding addresses using Google Maps, OpenStreetMap, and ArcGIS (5,14).

### Whole genome sequencing

Whole genome sequencing was conducted on 1,671 archived DNA samples with sufficient amounts of DNA (>0.05 ng/ul) for analysis. DNA was originally prepared by crude extraction from liquid culture samples and has been previously described (15). Libraries were prepared for sequencing using Nextera XT kit to obtain 2×150 base pair fragments for paired end sequencing, which was done using Illumina NextSeq 500 platform (15). Sequence assembly and analysis were performed using the MTBseq pipeline, which incorporates several open-source programs (including BWA, Samtools, and GATKv3) to automate steps involved in sample-specific and comparative analysis (15). Reads were mapped to the *M. tuberculosis* H37Rv reference genome (GenBank ID: NC_000962.3) (15). Variant calling was performed using default thresholds for coverage and quality (15). Phylogenetically informative single nucleotide polymorphisms (SNPs) were identified from existing literature. Annotations of variants associated with antibiotic resistance were based on a built in list of known mutations (16,17). Drug resistance summaries were generated for each sample to provide genotypic resistance prediction. A cluster detection algorithm was used to identify closely related strains within each lineage (1–4) using a distance threshold of ≤ 5 pairwise SNPs to establish bacterial genetic relatedness representing recent transmission (8).

### Spatial analysis

Our main analysis included participants residing in the greater Gaborone area with both WGS and geospatial data for home location available. We excluded participants with evidence of possible mixed strain infection (n = 29) (18), which were detected using a method based on a binomial test procedure (19). The subset of participants in outbreaks (large genotypic groups of 10 or more individuals) were the main focus of the analysis. We included participants with remote infection (ungrouped strains) as a comparison group to represent underlying population density

We conducted a preliminary analysis to compare the geographic distribution of participants’ residences with WGS data available to participants overall, to rule out false impressions of spatial clustering due to geographic sampling bias. We estimated the geographic median center point and standard deviational ellipse (directional distribution at 2 standard deviations) for both sets of participants. The median center is a measure of central tendency that is robust to outliers and minimizes the distance from the central location to all other points being analyzed. The standard deviational ellipse encompasses the majority of points along both the X axis and the Y axis, providing a representation of the geographic range and directional orientation of the points being analyzed. We then used these tools to characterize the geographic distribution of participants belonging to each outbreak, as well as those with remote infection. We used a Monte Carlo test of spatial segregation to measure geographic variation among the different outbreak groups (20). We generated 999 random permutations of group labels associated with each geographic point and compared results from the simulations to the observed geographic data to assess for significant differences.

We generated kernel density maps to visualize areas where participants were spatially aggregated. The kernel density method provides an estimate of spatial concentration in terms of points per unit area (16). It uses a moving window method with a weighting scheme (at each location, points closer to the center of the window are given a greater weight) and generates a smoothed map that displays areas of potential geospatial clustering (16). We generated maps for each genotypic cluster group and for ungrouped strains using a 1 km buffer window. For the sake of visual display, density is shown on a different scale for ungrouped participants (up to 40 per sq km) than for participants in the genotypic cluster groups (up to 5 per sq km) due to differences in size of the datasets.

We used spatial *K*-function analysis to estimate spatial clustering among participants in each outbreak (16). We then compared relative clustering over a range of distances (0–8000 meters) by estimating the difference in *K*-functions between participants in each outbreak group, and participants with remote infection (16,17). This method helps to identify spatial patterns after accounting for baseline clustering due to underlying population density (16,17). We used random permutations to obtain confidence bands for *K*-function differences. We generated plots with distance along the X-axis and *K*-function estimates along the Y-axis, and examined the shape and behavior of the *K*-function for interpretation (16). We assessed lines falling above or below the horizontal zero line to detect greater or lesser clustering in one group compared to the other, and lines falling outside the upper or lower confidence bands to detect statistically significant differences.

We then calculated SNP distances between each pair of participants in the same genotypic cluster group to assess whether relationships between geographic and genetic difference varied by group. We generated boxplots to display SNP distance summaries by group. Similarly, we calculated pairwise geographic distances (km) for participants within each group. We plotted pairwise geographic distance against pairwise SNP distance and tested for correlation between the two using Spearman’s *rho*.

We investigated possible spatial-temporal trends by measuring the geographic distance between the homes of the first participant diagnosed with TB in each genotypic cluster group, and each participant subsequently diagnosed within these genotypically defined cluster groups. We plotted date of diagnosis against geographic distance to visualize possible patterns.

Initial mapping and descriptive spatial analysis including median center, directional distribution, and kernel density estimation were performed using ArcGIS (version 10.7.1) (23) Additional analysis and data visualization were performed using R Statistical Software (version 4.1.2) (19). Pairwise geographic distances were calculated using the fields package (20) and pairwise SNP distances were calculated using the ape package (26). We used splancs (22) for *K*-function analysis. Boxplots and scatter plots were displayed using ggplot2 (23) and egg (29) with the viridis (25) color palette.

### Phylogenetic analysis

We generated a maximum likelihood phylogenetic tree using IQ-TREE (version 1.6.12) (31) to represent genetic relationships among Mtb strains. We specified an HKY substitution model, which allows for unequal base frequencies and unequal transition rates, and included an ascertainment bias correction (32). To construct the tree, we expanded our data set to include all participants with Mtb strains belonging to Lineage 4, after excluding isolates with evidence of possible mixed infection. For the phylogenetic tree presentation, we used a midpoint rooting approach. We highlighted the location of the main genotypic cluster groups in our analysis within the tree. Branches were expanded vertically from the node representing the estimated most recent common ancestor for each group to allow detailed visualization. We then projected phylogenetic trees on to maps for each of the groups, displaying the location of each Mtb isolate in the tree with a link drawn to its corresponding geographic location. We used the R packages ape (26), ggtree (33), phytools (34), rgdal (35), mapdata (36), and prettymapr (37) for tree annotation and visualization.

### Ethical considerations

The Kopanyo Study was approved by the U.S. Center for Disease Control and Prevention (CDC) Institutional Review Board (IRB) (#6291), the Health Research and Development Committee of the Botswana Ministry of Health and Wellness, and the University of Pennsylvania IRBs. We received written informed consent from all participants. Coordinates were mapped in low resolution to prevent identification.

## Results

A total of 1449 participants with culture-confirmed TB and primary residence in Greater Gaborone were enrolled in the original study. Of these, 946 (65%) had both WGS and home coordinates available. We noted participants with WGS data to be geographically representative of participants overall, based on comparison of geographic median center points and standard deviational ellipse. We excluded 29 participants with evidence of possible mixed strain infections. There were 431 participants that belonged to genotypic cluster groups of 2 to 19 people, including 62 participants belonging to five large genotypic cluster groups of 10 or more people. These groups (Group A – Group E) were considered outbreaks and were the focus of our current analysis, along with a control group of 486 participants that did not belong to a genotypic cluster (Table 1). Thus, the primary analysis was done on data from 548 study participants.

**Table 1.** Characteristics of study participants (N = 548) by outbreak group (≤ 5 SNP), Gaborone, Botswana, 2012-2016.

|  | Group A | Group B | Group C | Group D | Group E | Ungrouped |
| --- | --- | --- | --- | --- | --- | --- |
|  | (N = 19) | (N = 12) | (N = 11) | (N = 10) | (N = 10) | (N = 486) |
| Lineage | 4.1.1.3 | 4.1.1.3 | 4.1.2.1 | 4.1.1.3 | 4.3.4.1 |  |
| Age |  |  |  |  |  |  |
| Med | 29 | 35 | 33 | 31.5 | 39 | 35 |
| (IQR) | (24,40) | (28,40) | (31,42) | (30,37) | (34,42) | (28,42) |
| Gender |  |  |  |  |  |  |
| M | 11 (58%) | 9 (75%) | 9 (75%) | 3 (30%) | 10 (100%) | 254 (52%) |
| F | 8 (42%) | 3 (25%) | 3 (25%) | 7 (70%) | 0 (0%) | 232 (48%) |
| HIV |  |  |  |  |  |  |
| Pos | 9 (47%) | 6 (50%) | 9 (82%) | 5 (50%) | 5 (50%) | 308 (64%) |
| Neg | 10 (53%) | 6 (50%) | 2 (18%) | 5 (50%) | 4 (40%) | 162 (33%) |
| N/A | - | - | - | - | 1 (10%) | 16 (3%) |
| Income |  |  |  |  |  |  |
| Any | 15 (79%) | 5 (42%) | 8 (73%) | 4 (40%) | 8 (80%) | 360 (74%) |
| None | 4 (21 %) | 7 (58%) | 2 (18%) | 6 (60%) | 2 (20%) | 125 (25%) |
| N/A | - | - | 1 (9%) | - | - | 1 (<1%) |
| INH |  |  |  |  |  |  |
| S | 16 (84%) | 12 (100%) | 11 (100%) | 10 (100%) | 10 (100%) | 458 (94%) |
| R | 3 (16%) | 0 (0%) | 0 (0%) | 0 (0%) | 0 (0%) | 28 (6%) |
| RMP |  |  |  |  |  |  |
| S | 16 (84%) | 12 (100%) | 11 (100%) | 9 (90%) | 8 (80%) | 456 (94%) |
| R | 3 (16%) | 0 (0%) | 0 (0%) | 1 (10%) | 2 (20%) | 30 (6%) |

Note: INH, isoniazid; RMP, rifampicin.

Among participants with remote infections (ungrouped) the median age was 35 years (IQR: 28–42), just over half were male, one quarter reported no income, and 64% were diagnosed with TB-HIV coinfection (Table 1). The vast majority had Mtb that was susceptible to the first-line antibiotics isoniazid (INH; 94%) and rifampicin (RMP; 94%) based on genotypic prediction. Among participants in the five genotypic groups, median age ranged from 29 years (Group A) to 39 years (Group E) (Table 1). Participants in Group E were exclusively male, while Group D was the only genotypic group that had a greater proportion of female participants (70%). Group C was the only group with a majority of participants diagnosed with TB-HIV coinfection (9 of 11; 82%). The percentage of participants reporting no income ranged from 18% in Group C to 60% in Group D. Three participants in Group A had multidrug-resistant (MDR) TB with predicted resistance to both INH and RMP.

The maximum likelihood phylogenetic tree for Lineage 4 is shown in Figure 1. The location of isolates in each genotypic cluster group is highlighted, with a different color corresponding to each group. Group A, Group B, and Group D all belonged to sublineage 4.1.1.3 and were located near each other in the tree, while Group C and Group E belonged to sublineage 4.1.2.1 and 4.3.4.1 respectively and were located at greater genetic distances from the other groups.

**Figure 1.**
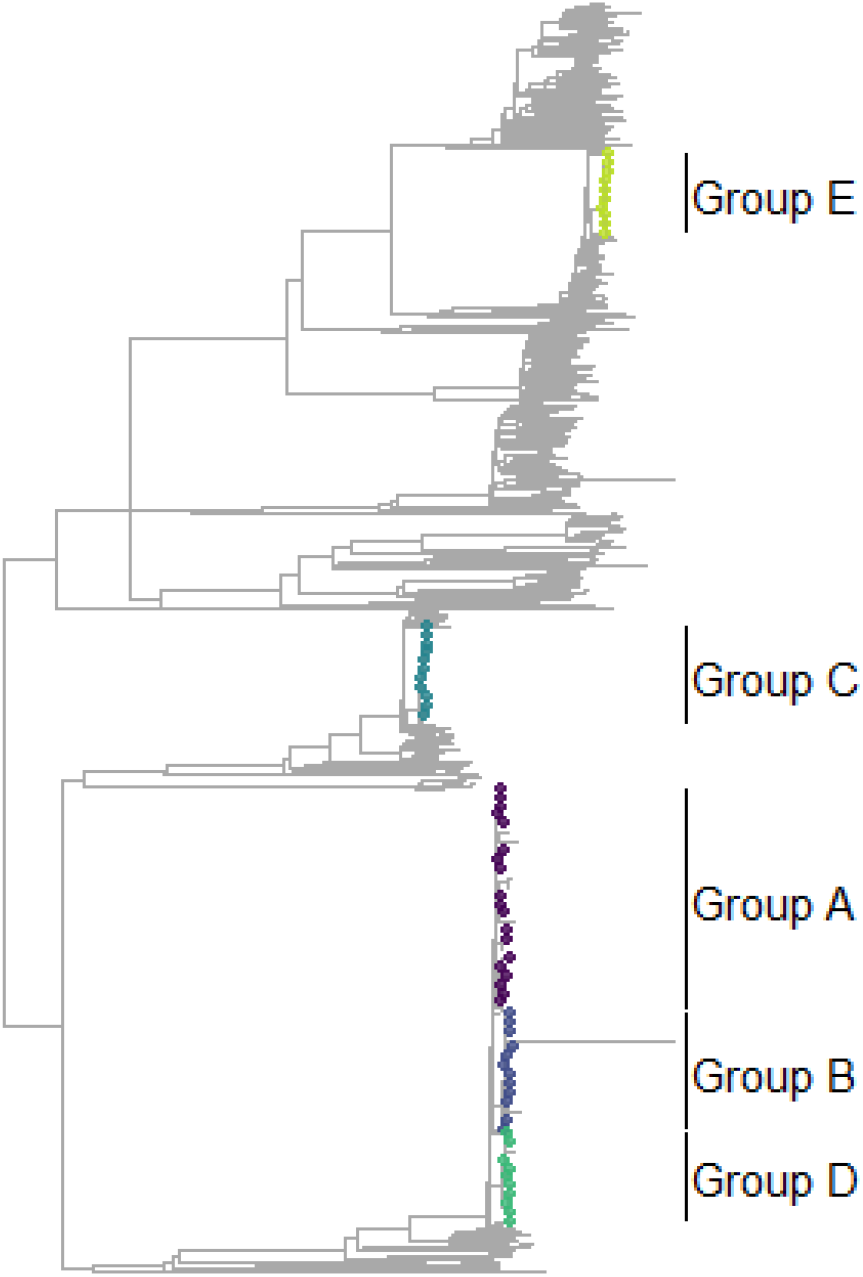
Phylogenetic tree for lineage 4 with location highlighted and expanded for selected genotypic cluster groups (≤ 5 SNP), Gaborone, Botswana, 2012–2016. Phylogenetic tree representation for all participants with *M. tuberculosis* strains belonging to Lineage 4. The location of isolates in each genotypic cluster group is highlighted. Branches within each of the groups were expanded for visualization.

Maps displaying kernel density estimation, median center points, and directional distributions for each outbreak group and for participants with remote infections are shown in Figure 2. We found evidence of distinct center points for each outbreak as demonstrated by spatial variation in geographic median center points for each group (Figure 3). The outbreak groups displayed significant spatial segregation (*p* = 0.038) indicating geographic variation among participants in different groups based on location of residence (Figure 2). Participants with ungrouped strains had residences widely spread across the study area (Figure 2, Table 2). The distance between the center points of ungrouped strains and each of the genotypic groups ranged from 458 meters for Group A to nearly 4,979 meters for Group D (Figure 3, Table 2). Participants in Group C had an elongated (12.5 km) east-west distribution, while Group B (9 km) and Group D (8 km) both had a more compact north-south spread (Figure 2, Table 2). Areas of spatial clustering of residences among participants within each group were visually apparent from kernel density estimation, and were especially pronounced in the south-central part of the map for Group B and Group D (Figure 2).

**Table 2.** Spatial summary for each *M. tuberculosis* outbreak group (≤ 5 SNP) by distance rank from reference, Gaborone, Botswana, 2012-2016.

| Distance Rank | Group | Median Center Distance (m) | X Span (m) | Y Span (m) | Rotation (degrees) |
| --- | --- | --- | --- | --- | --- |
| 1 | Group A | 458 | 9120 | 10263 | 28 |
| 2 | Group E | 1633 | 9597 | 5983 | 134 |
| 3 | Group C | 3143 | 12431 | 4768 | 111 |
| 4 | Group B | 3788 | 6251 | 9077 | 15 |
| 5 | Group D | 4979 | 5946 | 8197 | 29 |
| Reference | Ungrouped | Reference | 11786 | 9507 | 103 |

**Figure 2.**
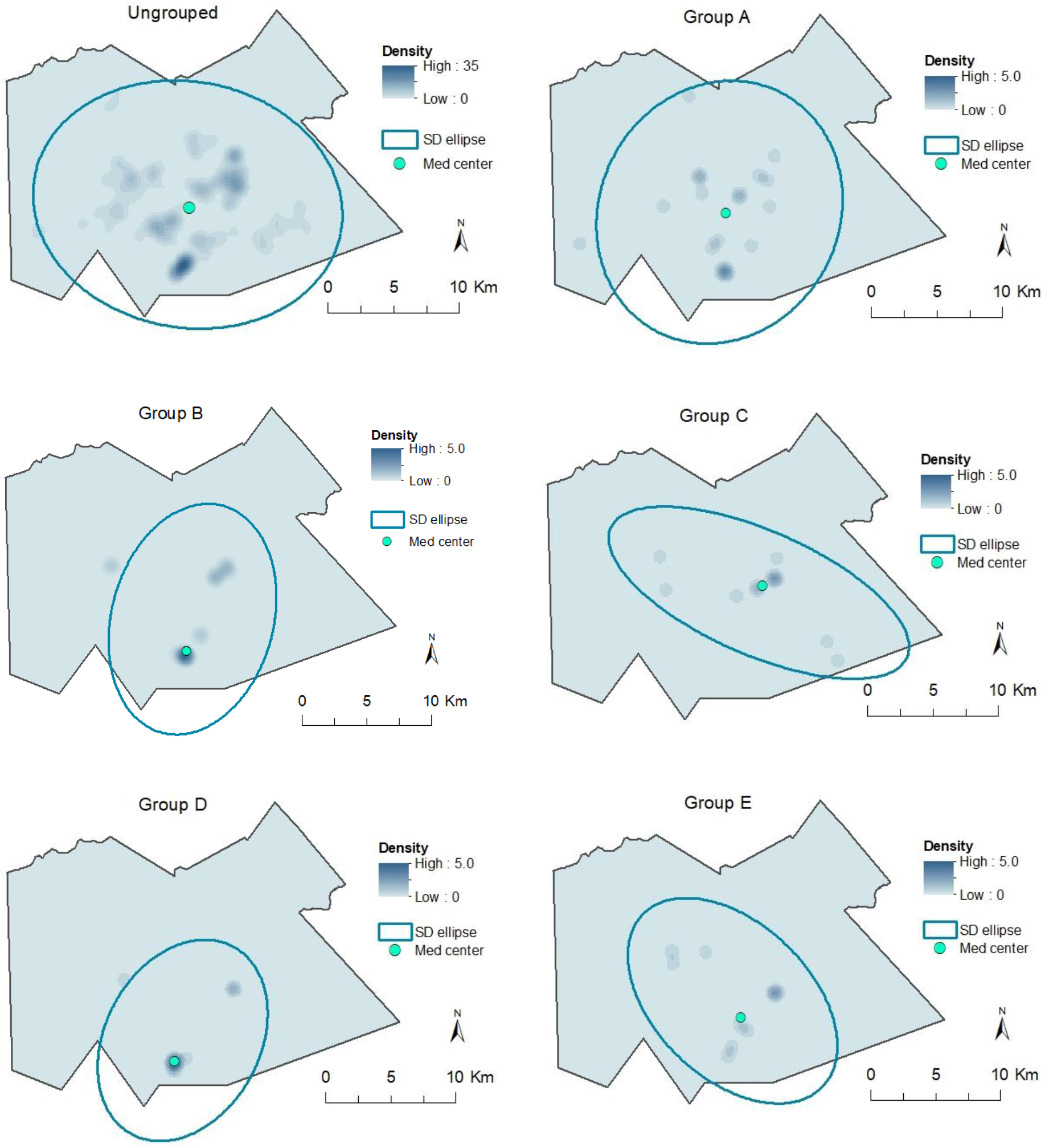
Kernel density map, median center point, and directional distribution for each *M. tuberculosis* genotypic cluster group (≤ 5 SNP) and ungrouped strains, Gaborone, Botswana, 2012-2016. Kernel density maps and standard deviational ellipse based on location of primary residence for ungrouped participants and participants in each genotypic cluster group. The blue ovals represent the area within the standard deviational ellipse, representing the geographic range (km) and directional orientation of participant locations within each group. Density is shown on a different scale for ungrouped participants (up to 40 per sq km) than for participants in the genotypic cluster groups (up to 5 per sq km) due to differences in size of the datasets.

**Figure 3.**
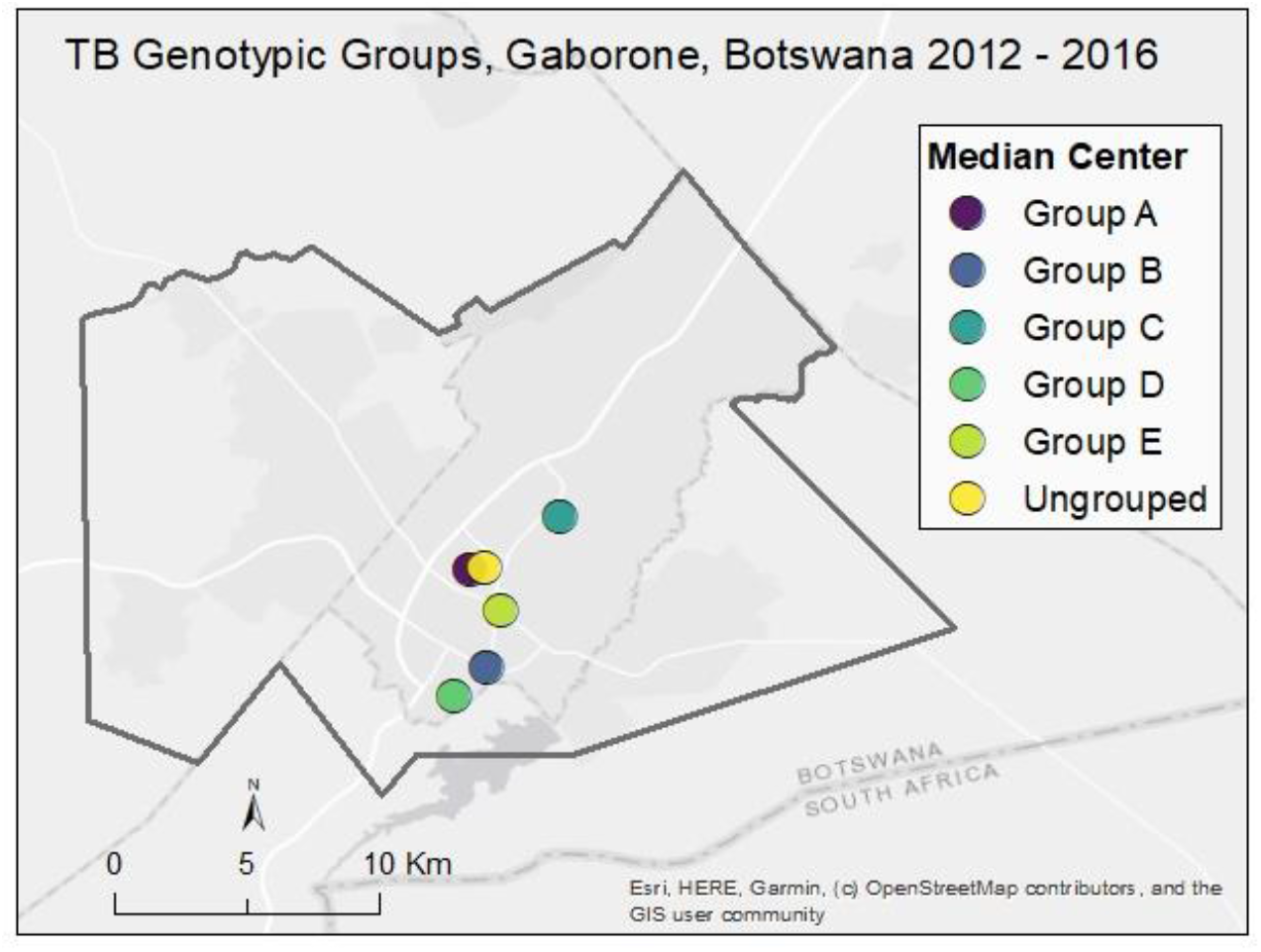
Median center point for each *M. tuberculosis* genotypic group (≤ 5 SNP) and ungrouped strains, Gaborone, Botswana, 2012–2016. Location of geographic median center point for ungrouped participants and participants in each genotypic cluster group. The median center represents a centralized geographic location that is estimated by minimizing the distance to all other locations being analyzed (in this case, locations of participants in each group).

Results from the *K*-function analysis are displayed in Figure 4. Differences in *K*-functions indicated participants in Group B and Group D had greater spatial clustering than participants with ungrouped strains, falling above the upper 95% confidence band up to a distance of approximately 4 km. Group A and Group E had similar clustering to ungrouped strains, while Group C had greater clustering but not consistently significant.

**Figure 4.**
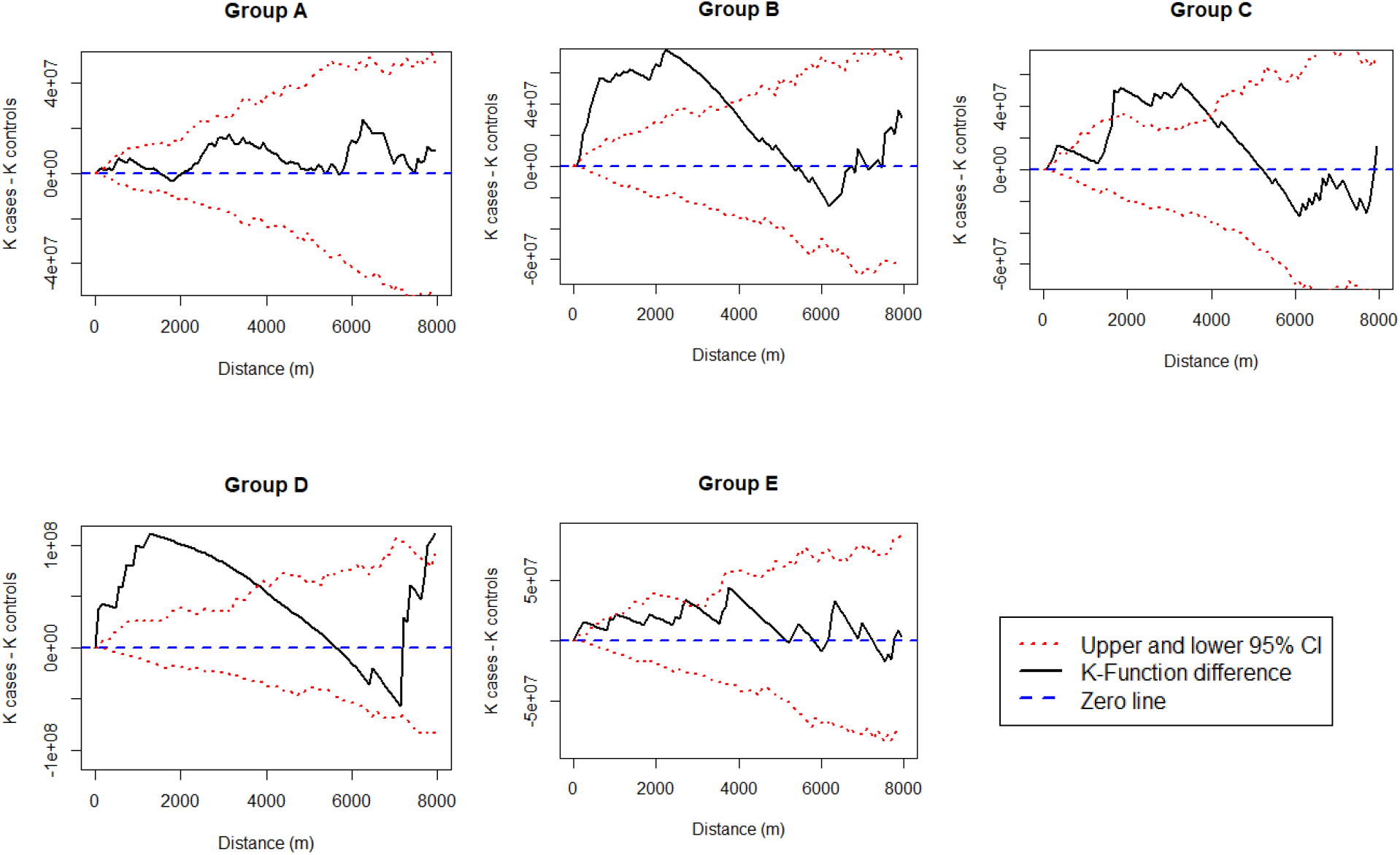
*K*-function differences for each *M. tuberculosis* genotypic cluster group (≤ 5 SNP) compared to ungrouped strains, Gaborone, Botswana, 2012–2016. Differences in spatial K-functions for each *M. tuberculosis* genotypic cluster group (≤ 5 SNP) compared to ungrouped strains. Lines falling above or below the horizontal zero line indicate greater or lesser spatial clustering in one group compared to the other. Lines falling above or below the upper or lower confidence bands indicate statistically significant differences.

Time between diagnoses by geographic distance from the first participant detected in each group is shown in Figure 5. Temporal trends in geographic distance between the first and subsequent cases varied by group. Group C had an overall pattern of increasing distance with increasing time, while for Group D all subsequent cases were relatively near to the first participant diagnosed. In Group B the first detected case was located at a relatively large but equal distance from all subsequently diagnosed participants.

**Figure 5.**
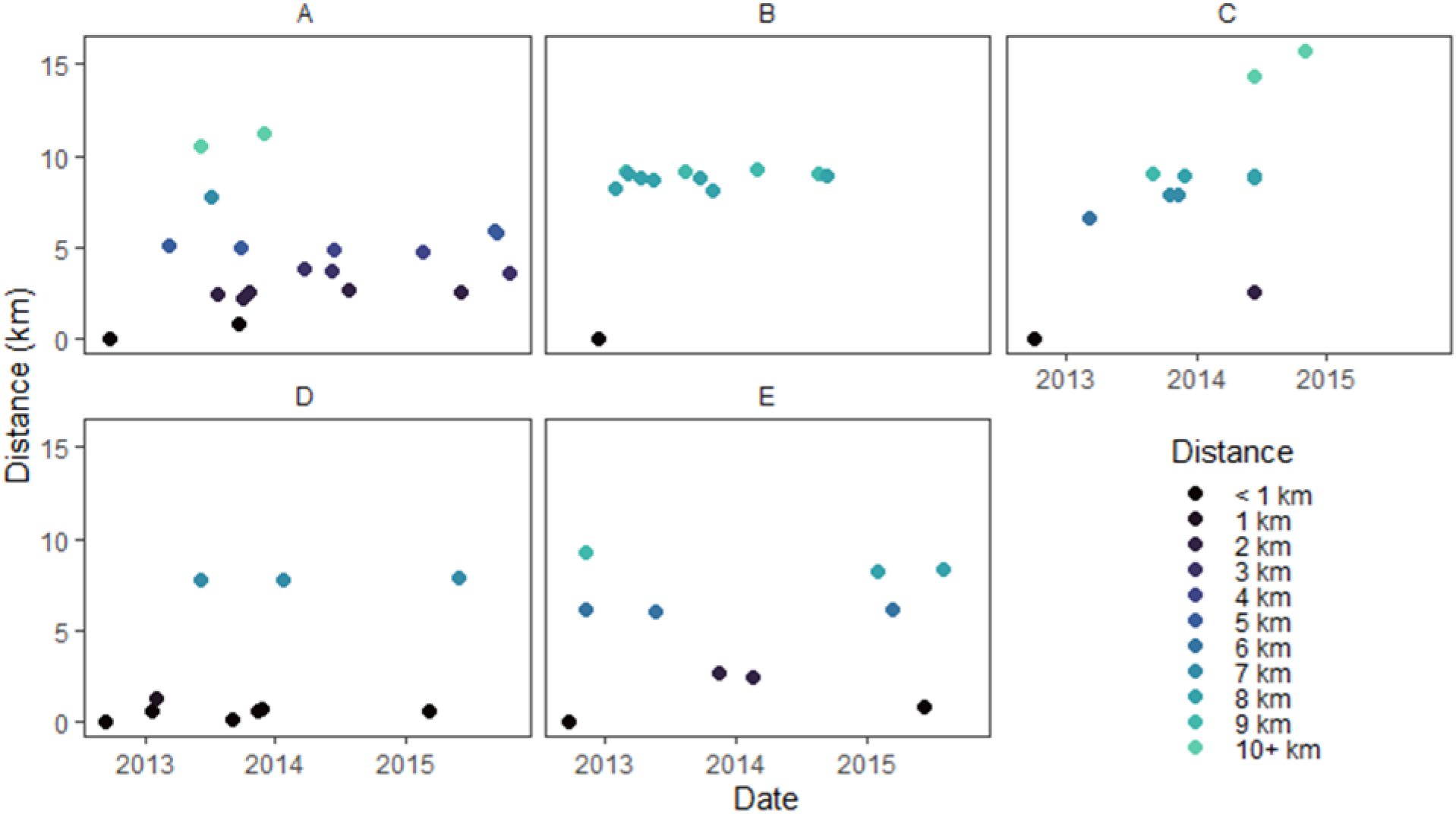
Incident TB by geographic distance from first participant diagnosed by genotypic cluster group (≤ 5 SNP), Gaborone, Botswana, 2012–2016. Plots representing each participant by date of TB diagnosis and by geographic distance (km, based on participant’s primary residence) from the first participant (shown in each plot at a distance of 0) diagnosed in each genotypic cluster group.

Median within-group SNP distance was below 5 SNPs for all groups except Group A, which had a median of 7 (Figure 6). Group A also had higher variability in pairwise SNP distances compared to other groups. We observed a low level of positive correlation between geographic and SNP pairwise distances overall (*rho* = 0.1; *p* = 0.06). However, this varied by group, with Group A and Group E displaying low to modest positive correlation (*rho* =0.26; *p* = 0.001, and 0.3; *p* = 0.045, respectively) while the correlation for Group C was negative (*rho* = - 0.33; *p* = 0.015).

**Figure 6.**
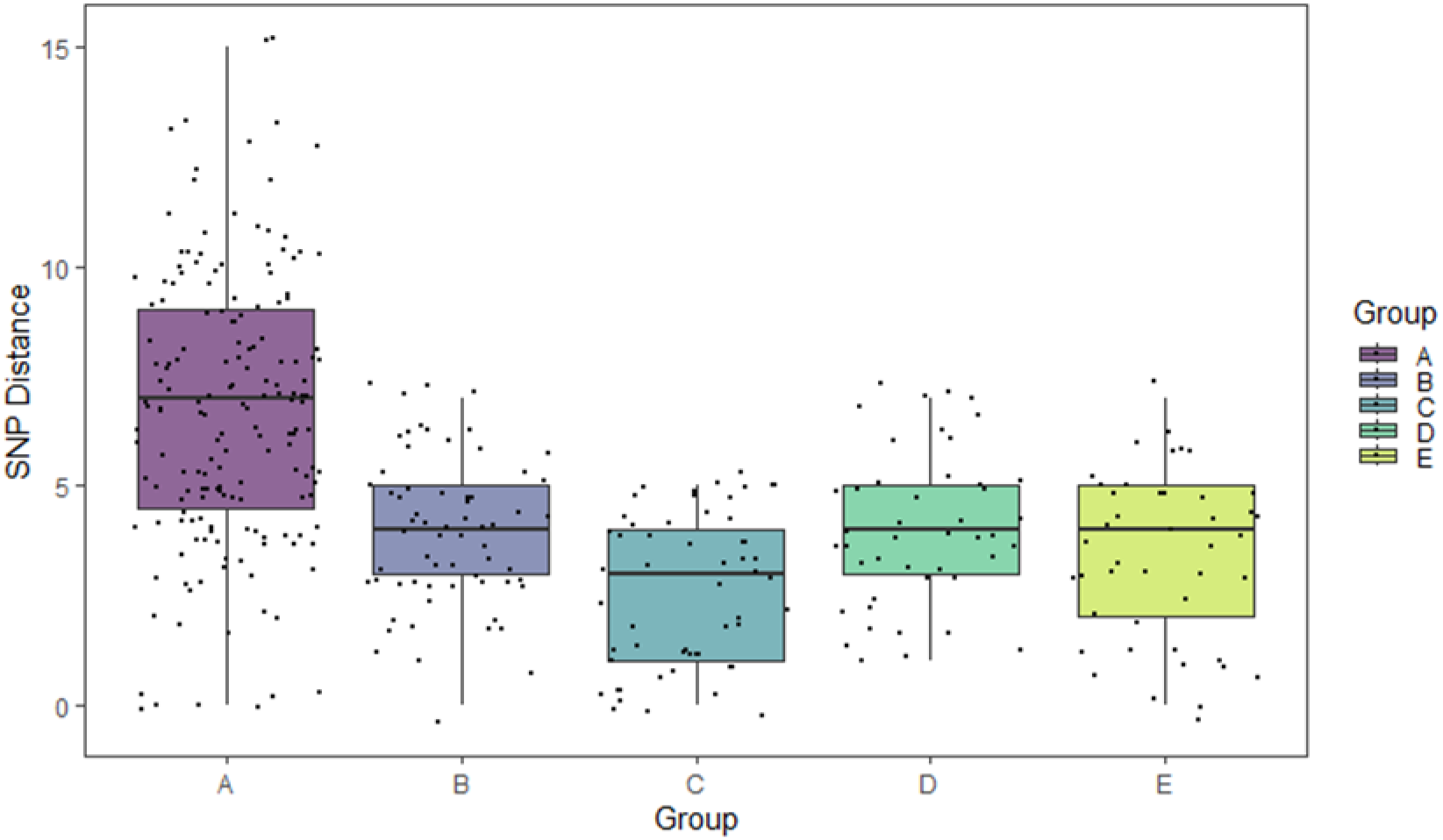
Pairwise SNP distances by genotypic cluster group (≤ 5 SNP), Gaborone, Botswana, 2012–2016. Box plots with individual data points superimposed displaying SNP distance summaries by group. Median within-group SNP distance was below 5 SNPs for all groups except Group A, which had a median of 7 SNPs.

**Figure 7.**
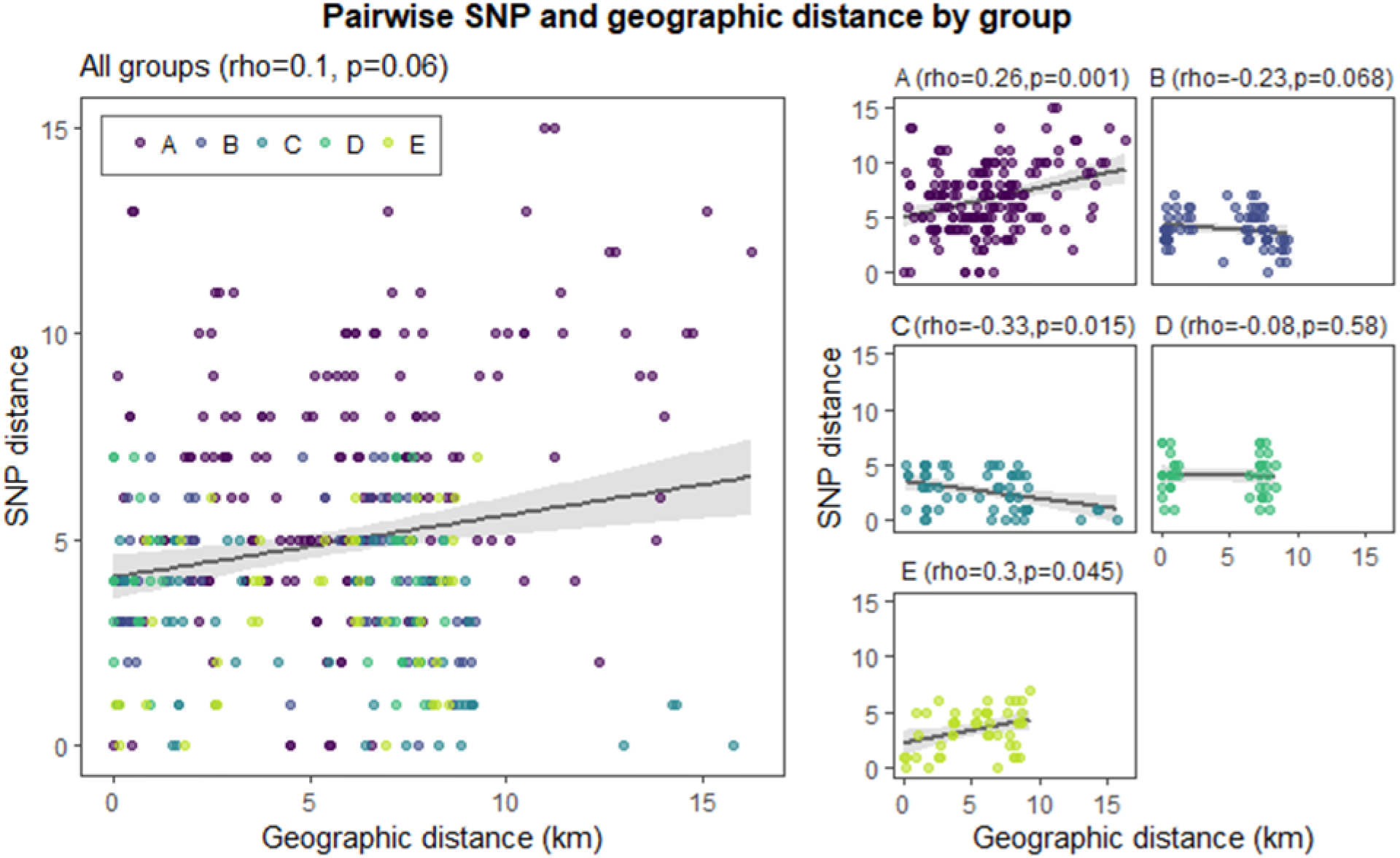
Correlation between pairwise SNP distance and pairwise geographic distance by genotypic cluster group (≤ 5 SNP), Gaborone, Botswana, 2012-2016. Comparisons of pairwise geographic and SNP distances. Points represent measurements for within-group pairs. There was low positive correlation between pairwise geographic and SNP distances overall (Spearman’s *rho* = 0.1; *p* = 0.06).

Phylogenetic tree displays linked to spatial maps are shown in Figure 8. Heterogenous genotypic and geographic patterns were seen in the different groups. In Group E, closely related Mtb isolates were also generally located closer in space, and separate areas of potential geographic clustering were visible. In Group D, the majority of isolates appeared to aggregate in a single geographic cluster, regardless of within-group genetic relatedness. A similar pattern was seen in Group B with two potential spatial clusters. In Group A and Group C, closely related isolates were generally dispersed more broadly over the geographic area. Distinct subclusters of spatially and phylogenetically linked cases could be identified in all groups.

**Figure 8.**
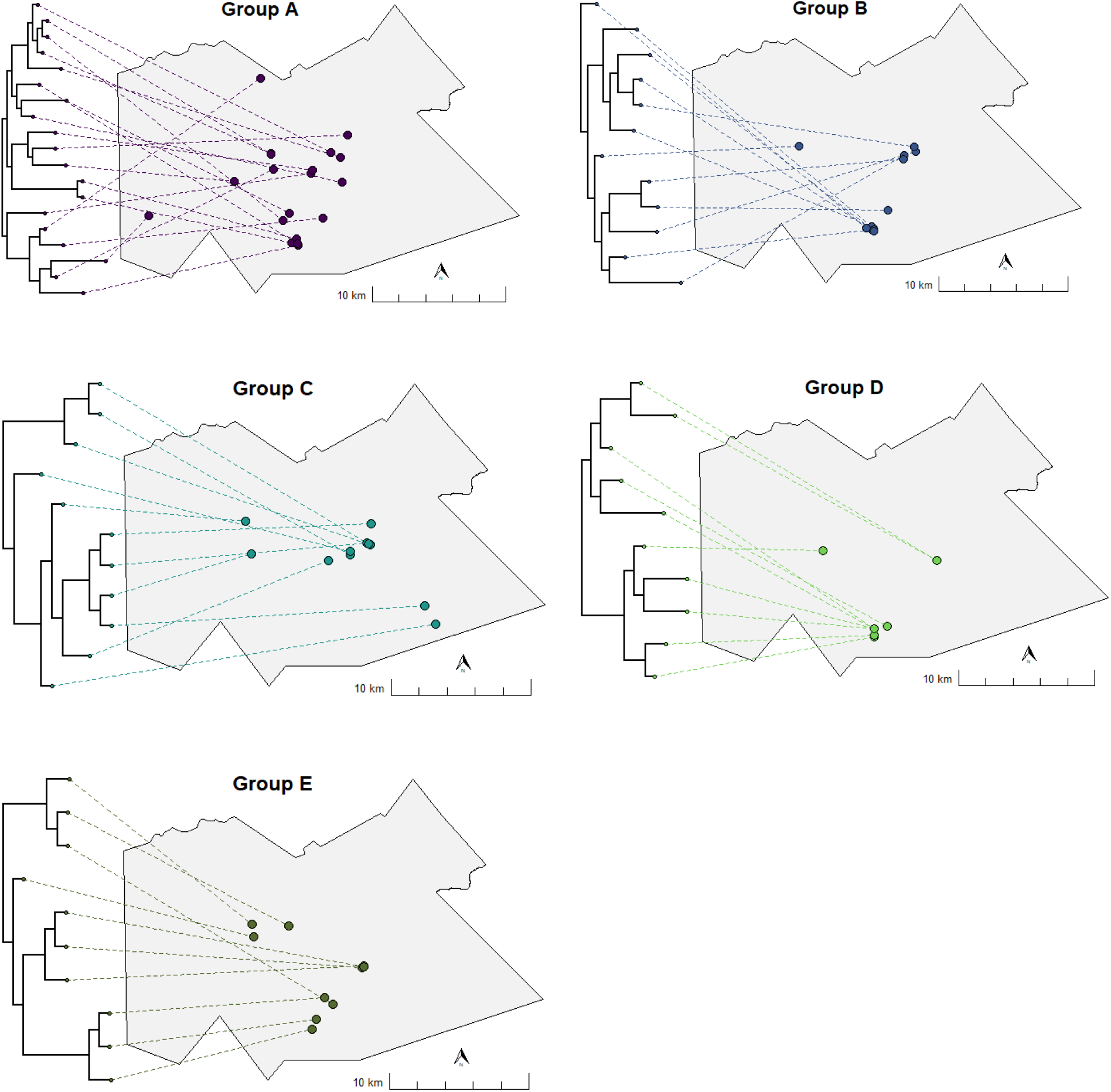
Representation of phylogenetic trees projected on to maps for each genotypic cluster group (≤ 5 SNP), Gaborone, Botswana, 2012-2016. Representation of phylogenetic tree data projected on to geographic maps for each group. The location of each *M. tuberculosis* isolate in the tree is displayed with a link drawn to its corresponding geographic location. Tree tips on the same bifurcating branches represent the most closely related isolates.

## Discussion

In our analysis, genotypic cluster groups of closely related Mtb strains displayed distinct geospatial characteristics. Spatial clustering among participants in two of the genotypic cluster groups may indicate localized areas of recent transmission. Genotypic cluster groups that were less spatially clustered may be transmitting more broadly through the study area. For example, strains belonging to Group A had greater genetic and spatial heterogeneity, suggesting a more advanced stage in the transmission trajectory compared to other groups.

Temporally, the geographic distance between the first diagnosed and subsequently diagnosed cases varied by group. Interestingly, the first diagnosed case in Group B was located at a relatively large but equal distance from all subsequently diagnosed participants. Further probing of spatial data could reveal whether these participants were spatially clustered, potentially indicating an outside introduction of TB that led to a localized transmission cluster. However, this observation could also be related to the timing of sampling, missed cases, or incomplete spatial data. In contrast to focusing on the location of the first detect case in a genotypic cluster, a location-based approach around the most recently diagnosed case has been suggested as a high yield approach for active case finding (38).

Our results support findings from our prior analysis, which found evidence of localized transmission by detecting spatial clustering of genotypic groups identified using MIRU-VNTR (5). While the overall areas of spatial aggregation were similar, our current analysis builds on previous work by incorporating higher resolution genomic sequencing data to detect finer-scale spatial patterns and describe the geographic distribution of distinct genotypic groups.

Our results also align with recent studies combining spatial and WGS data to study TB transmission in several other high burden settings, including China (33,34), Ghana (41), and along the Thai-Myanmar border (36). Observed spatial patterns among related Mtb strains have included local and regional distributions of genotypic cluster groups (39,41) and lineages (42), and associations between residential proximity and genetic similarity (33,34) In contrast, a study in China found the majority of genotypic groups included participants from separate geographic districts (43). However, that study differed from ours because it specifically analyzed MDR-TB, and included a majority of participants (70%) that had migrated from other provinces (43).

Phylogenetic trees and geographic maps are often presented as complementary but separate displays of data. We generated phylogenetic trees linked to spatial maps that produced a high-resolution display for each genotypic cluster that could guide public health activities. For example, potential sub-groupings of closely related strains within genotypic cluster groups could be linked to their corresponding geographic locations, which could help identify high risk areas for targeted interventions, including active case finding for early diagnosis and linkage to care, contact investigations, and TB preventive therapy.

One limitation to our analysis was that WGS and geospatial data were not available for all study participants. While the samples included in this study were found to be geographically representative of the larger study population overall, smaller scale spatial patterns could have been affected. We also excluded possible mixed strain infections, which could have led to missed detection of Mtb sequences belonging to the same genotypic groups (18,44). In addition, we did not include data on epidemiological links among social contacts or potential community sites of transmission. Additional WGS and epidemiologic data would contribute to a more complete reconstruction and spatial representation of transmission networks (45).

## Conclusion

The integration of genomic and geospatial data is a promising approach for studying TB transmission in high burden settings. We used this approach to identify heterogeneity between multiple Mtb transmission chains. We identified geographically clustered strains of Mtb representing localized areas of recent transmission. Used in near real time, this could potentially help TB prevention programs identify emerging outbreaks. While barriers remain, significant progress has been made toward increasing capacity for genomic technologies in low and middle-income countries (43,44). Integrated genomic and geospatial approaches could help with planning and mobilizing interventions to interrupt ongoing transmission (43,44).

## Supporting information

Research reporting checklist (STROME-ID)

## Data Availability

Data produced in the present study are available upon reasonable request to the authors. Sequencing data for M. tuberculosis genes will be deposited into Genbank (http://www.ncbi.nlm.nih.gov/genbank/). To protect confidentiality, GPS coordinates will not be included in the shared dataset.

