## Supplementary material for "High-resolution characterization of recent tuberculosis transmission in Botswana using geospatial and genomic data – the Kopanyo Study": Research reporting checklist (STROME-ID)

Table The STROBE checklist and additional STROME-ID items

| | Item number | STROBE items | STROME-ID items |
|---|---|---|---|
| **Title and abstract** | | | |
| Introduction | 1 | (a) Indicate the study's design with a commonly used term in the title or the abstract<br>(b) Provide in the abstract an informative and balanced summary of what was done and what was found | STROME-ID 1.1: the term molecular epidemiology should be applied to the study in the title or abstract and the keywords when molecular and epidemiological methods contribute substantially to the study |
| Background rationale | 2 | Explain the scientific background and rationale for the investigation being reported | STROME-ID 2.1: provide background information about the pathogen population and the distribution of pathogen strains within the host population at risk |
| Objectives | 3 | State specific objectives, including any prespecified hypotheses | STROME-ID 3.1: state the epidemiological objectives of using molecular typing |
| **Methods** | | | |
| Study design | 4 | Present key elements of study design early in the paper | .. |
| Molecular terminology | | .. | STROME-ID 4.1: define or cite definitions for key molecular terms used within the study (eg, strain, isolate, and clone) |
| Molecular markers | | .. | STROME-ID 4.2: clearly define the molecular markers that were used with a standard nomenclature |

| | Item number | STROBE items | STROME-ID items |
|---|---|---|---|
| Infectious disease case definition | | .. | STROME-ID 4.3: clearly state the infectious-disease case definitions |
| Laboratory methodology | | .. | STROME-ID 4.4: describe sample collection and laboratory methods, including any methods used to minimise and measure cross-contamination, and give the criteria used to interpret strain classification |
| Setting | 5 | Describe the setting, locations, and relevant dates, including periods of recruitment, exposure, follow-up, and data collection | STROME-ID 5.1: clearly state the timeframe of the study; consider and appropriately reference the molecular clock of markers if known, and the natural history of the infection |
| Participants | 6 | (a) *Cohort study*—give the eligibility criteria, and the sources and methods of selection of participants. Describe methods of follow-up *Case-control study*—give the eligibility criteria, and the sources and methods of case ascertainment and control selection. Give the rationale for the choice of cases and controls *Cross-sectional study*—give the eligibility criteria, and the sources and methods of selection of participants (b) *Cohort study*—for matched studies, give matching criteria and number of exposed and unexposed *Case-control study*—for matched studies, give matching criteria and the number of controls per case | STROME-ID 6.1: state the source of participants and clinical specimens, and clearly describe sampling frame and strategy |

| | Item number | STROBE items | STROME-ID items |
|---|---|---|---|
| Variables | 7 | Clearly define all outcomes, exposures, predictors, potential confounders, and effect modifiers. Give diagnostic criteria, if applicable | .. |
| Data sources/measurement | 8 * | For each variable of interest give sources of data and details of methods of assessment (measurement). Describe comparability of assessment methods if there is more than one group | .. |
| Multiple-strain infections | | .. | STROME-ID 8.1: describe any methods used to detect multiple-strain infections and measure their effect on the study findings |
| Bias | 9 | Describe any efforts to address potential sources of bias | STROME-ID 9.1: describe any efforts made to address discovery or ascertainment bias |
| Study size | 10 | Explain how the study size was arrived at | STROME-ID 10.1: describe any unique restrictions placed on the study sample size |
| Quantitative variables | 11 | Explain how quantitative variables were handled in the analyses. If applicable, describe which groupings were chosen, and why | .. |

| | Item number | STROBE items | STROME-ID items |
|---|---|---|---|
| Statistical methods | 12 | (a) Describe all statistical methods, including those used to control for confounding<br>(b) Describe any methods used to examine subgroups and interactions<br>(c) Explain how missing data were addressed<br>(d) *Cohort study*—if applicable, explain how loss to follow-up was addressed<br>*Case-control study*—if applicable, explain how matching of cases and controls was addressed<br>*Cross-sectional study*—if applicable, describe analytical methods taking account of sampling strategy<br>(e) Describe any sensitivity analyses | STROME-ID 12.1: state how the study took account of the non-independence of sample data, if appropriate<br>STROME-ID 12.2: state how the study dealt with missing data |
| **Results** | | | |
| Participants | 13* | (a) Report the numbers of individuals at each stage of the study (eg, numbers potentially eligible, examined for eligibility, confirmed eligible, included in the study, completing follow-up, and analysed)<br>(b) Give reasons for non-participation at each stage<br>(c) Consider use of a flow diagram | STROME-ID 13.1: Report numbers of participants and samples at each stage of the study, including the number of samples obtained, the number typed, and the number yielding data<br>STROME-ID 13.2: if the study investigates groups of genetically indistinguishable pathogens (molecular clusters), state the sampling fraction, the distribution of cluster sizes, and the study population turnover, if known |

| | Item number | STROBE items | STROME-ID items |
|---|---|---|---|
| Descriptive data | 14[*] | (a) Give characteristics of study participants (eg, demographic, clinical, social) and information on exposures and potential confounders<br>b) Indicate the number of participants with missing data for each variable of interest<br>(c) *Cohort study*—summarise follow-up time (eg, average and total amount) | STROME-ID 14.1: give information by strain type if appropriate, with use of standardised nomenclature |
| Outcome data | 15[*] | *Cohort study*—report numbers of outcome events or summary measures over time<br>*Case-control study*—report numbers in each exposure category, or summary measures of exposure<br>*Cross-sectional study*—report numbers of outcome events or summary measures | .. |
| Main results | 16 | (a) Give unadjusted estimates and, if applicable, confounder-adjusted estimates and their precision (eg, 95% confidence interval). Make clear which confounders were adjusted for and why they were included<br>(b) Report category boundaries when continuous variables were categorised<br>(c) If relevant, consider translating estimates of relative risk into absolute risk for a meaningful time period | STROME-ID 16.1: consider showing molecular relatedness of strain types by means of a dendrogram or phylogenetic tree |
| Other analyses | 17 | Report other analyses done (eg, analyses of subgroups and interactions, and sensitivity analyses) | .. |

| | Item number | STROBE items | STROME-ID items |
|---|---|---|---|
| **Discussion** | | | |
| Key results | 18 | Summarise key results with reference to study objectives | .. |
| Limitations | 19 | Discuss limitations of the study, taking into account sources of potential bias or imprecision. Discuss both direction and magnitude of any potential bias | STROME-ID 19.1: consider alternative explanations for findings when transmission chains are being investigated, and report the consistency between molecular and epidemiological evidence |
| Interpretation | 20 | Give a cautious overall interpretation of results considering objectives, limitations, multiplicity of analyses, results from similar studies, and other relevant evidence | .. |
| Generalisability | 21 | Discuss the generalisability (external validity) of the study results | .. |
| **Other information** | | | |
| Funding | 22 | Give the source of funding and the role of the funders for the present study and, if applicable, for the original study on which the present article is based | .. |
| Ethics | 23 | .. | STROME-ID 23.1: report any ethical considerations with specific implications for infectious-disease molecular epidemiology |

STROBE=Strengthening the Reporting of Observational Studies in Epidemiology. STROME-ID=Strengthening the Reporting of Molecular Epidemiology for Infectious Diseases.

* Give such information separately for cases and controls in case-control studies, and, if applicable, for exposed and unexposed groups in cohort and cross-sectional studies.
